# Brain structure of South African children born to mothers on dolutegravir versus efavirenz-based ART

**DOI:** 10.1101/2025.10.10.25329474

**Authors:** Layla E Bradford, Catherine J Wedderburn, Thokozile R Malaba, Helene Theunissen, Jessica E Ringshaw, Niall J Bourke, Steve CR Williams, Nengjie He, Lucy Read, Catriona Waitt, Helen Reynolds, Angela Colbers, Jim Read, Lauren Davel, Catherine Orrell, Miriam Taegtmeyer, Duolao Wang, Saye Khoo, Landon Myer, Kirsten A Donald

## Abstract

**Background:** The impact of in utero exposure to specific antiretroviral therapy (ART), particularly integrase strand transfer inhibitors, on early brain development remains poorly understood. We used magnetic resonance imaging (MRI) to compare brain structure in children who are HIV-exposed and uninfected (CHEU) with prenatal dolutegravir (DTG) versus efavirenz (EFV) exposure

**Methods:** DolPHIN-2 was a randomized trial of pregnant women initiating DTG-versus EFV-based ART in the third trimester. At 3-4 years of age, a subgroup of their children from the South African cohort, along with HIV-unexposed children (CHU), underwent T1-weighted MRI (DolPHIN-2 PLUS). Measurements of brain structure including volume, cortical thickness, and surface area were extracted using Freesurfer. Associations between ART /HIV exposure and child brain structure were examined using multiple linear regression.

**Results:** This analysis included 58 children (25 CHEU [13 DTG, 12 EFV] and 33 CHU, mean age 46.35 months, 51.7% male). Demographic characteristics were similar across groups. No significant differences in global or regional brain volumes, cortical thickness, or surface area were observed between DTG- and EFV-exposed children or between CHEU and CHU.

**Conclusion:** Among children exposed to ART in the third trimester, brain structural measures at 3–4 years were comparable between DTG and EFV exposure. CHEU also showed similar brain development to CHU. Further longitudinal studies with larger sample sizes are needed to assess the effects of specific ART in the context of new regimens and better maternal HIV disease control, on CHEU brain structure.

## Introduction

There is a growing population of children who are HIV-exposed and uninfected (CHEU), particularly in Sub-Saharan Africa^1,2^. CHEU are considered a vulnerable population, with evidence indicating an increased risk for adverse health outcomes including immune, growth and developmental alterations^3-7^. Neurodevelopmental differences including poorer language and motor outcomes have been noted in CHEU compared to children who are HIV-unexposed (CHU) from similar contexts^8-10^. This highlights a significant health burden and barrier to realizing the full developmental potential in this population.

One potential contributor to these neurodevelopmental differences is prenatal exposure to antiretroviral therapy (ART). While maternal ART usage during pregnancy is essential for preventing vertical HIV transmission and supporting maternal immune health, the potential neurotoxic effects on fetal brain development remain poorly understood^11^. Early life represents a critical and sensitive period of brain growth, and this lack of data and limits our ability to fully evaluate the long-term risks and benefits of current ART regimens used during pregnancy. There is some evidence suggesting an association between efavirenz (EFV)-based regimens and microcephaly as well as poorer neurodevelopmental outcomes, although findings are inconsistent^9,10,12-15^.

Neuroimaging, particularly magnetic resonance imaging (MRI), has the potential to offer valuable insight into the neurobiological mechanisms and contributing factors involved in the neurodevelopment of CHEU^16^. A South African neuroimaging study of HIV-exposed but uninfected (HEU) neonates exposed in utero to EFV-based ART found that those born to mothers who initiated ART before conception had larger caudate volumes than those whose mothers began treatment later in pregnancy, suggesting that longer ART duration may protect against subcortical brain alterations in CHEU^17^. This suggests ART, by improving maternal immune functioning through viral suppression, may offer neuroprotection to the developing fetus. Consistent with these findings, other studies have found associations between poor maternal immune status (low maternal CD4 counts and high viral loads) and smaller subcortical brain volumes, reinforcing the connection between maternal immune regulation and fetal brain development in the context of HIV^18,19^.

While there is a global effort to develop safer and more efficacious drugs for the treatment of HIV, pregnant women are often excluded from ART clinical trials due to safety concerns, resulting in limited data on the safety and long-term effects of in utero ART exposure. The DolPHIN-2 trial investigated dolutegravir (DTG), an integrase strand transfer inhibitor, in comparison to EFV, a non-nucleoside reverse transcriptase inhibitor, in pregnant women initiating ART in the third trimester^20^. Results showed that DTG achieved a more rapid and greater viral load suppression by the time of delivery compared to EFV^20^. However, no studies to date have examined the impact of prenatal DTG exposure on child brain structure.

In this study, we address this gap by comparing brain structure at 3–4 years of age in children exposed in utero to DTG versus EFV, using MRI. Secondary objectives included comparing CHEU and CHU brain structure and examining associations between maternal HIV disease severity and child brain development.

## Methods

### Participants

DolPHIN-2 PLUS is an observational infant-follow up sub-study of the DolPHIN-2 trial (NCT03249181) in pregnant women who initiated ART (DTG vs. EFV) in the third trimester^20^. Participants were recruited from the Gugulethu Midwife Obstetrics Unit, a primary prenatal healthcare facility located in Gugulethu, an informal peri-urban settlement within the Cape Town metropole with a predominantly isiXhosa-speaking population. Gugulethu, like many other informal settlements in South Africa, is impacted by a significant health burden caused by a high prevalence HIV. CHEU from DolPHIN-2 were invited to participate in DolPHIN-2 PLUS alongside a group of CHU, who were recruited postnatally using convenience sampling and were similar in age, sex and sociodemographic background. Overall, 58 children (25 CHEU and 33 CHU), born between 2018 and 2019, were included in this analysis aged between 39-54 months. Children were excluded if they had a known positive HIV test or MRI contraindication.

### Procedures

#### HIV/ART data

Study procedures in the pregnant mothers from DolPHIN-2 have been described previously^20^. In the third trimester, mothers in the DTG group received dolutegravir (50 mg) plus generic tenofovir disoproxil fumarate (300 mg) co-formulated with emtricitabine (200 mg) and mothers in the EFV group received a generic single fixed-combination pill of efavirenz (600 mg) with tenofovir and emtricitabine. Maternal viral load was measured at enrolment during the third trimester as well as within 14 days post-delivery. CD4 was measured at enrolment.

#### Sociodemographic variables

Data on sociodemographic characteristics of mothers were collected using a combination of tools, including the Edinburgh Postnatal Depression Scale (EPDS), Alcohol, Smoking and Substance Involvement Screening Test (ASSIST) and a study-designed socioeconomic questionnaire.

#### Neuroimaging acquisition

Magnetic resonance imaging was conducted during non-sedated natural sleep using a Siemens 3T Skyra system, housed at The Cape Universities Brain Imaging Centre, located in Cape Town, South Africa. T1-weighted (T1w) MEMPRAGE scans were conducted to obtain brain morphological measures including cortical thickness, volume and surface area. The center provided a child-friendly environment for pediatric neuroimaging and the research team were familiar with the imaging procedures and followed a protocol described previously^16^.

#### Neuroimaging processing

Visual quality inspection was conducted on all scans to assess for the presence of motion artifact. T1w images were processed using Freesurfer version 7.2.0^21,22^. The automated Freesurfer pipeline (recon-all command) was run via an inbuilt gear on Flywheel (https://flywheel.io), a medical imaging and AI platform. As an end-to-end tool for data management, curation, and computational analysis, Flywheel served as the host for centralized image processing. Global and regional subcortical brain volumes, cortical thickness, and surface area were extracted for analysis. This was computed according to the Desikan-Killiany atlas for cortical parcellation and an inbuilt probabilistic atlas for segmentation^23,24^. Based on prior research on HIV exposure and child brain outcomes, the regions of interest (ROIs) for this study were selected *a priori* and included total grey matter, total white matter, and several subcortical grey matter structures (thalamus, caudate, putamen, pallidum, hippocampus, and amygdala)^6,17-19^. An overview of the workflow for acquisition, processing, and analysis of structural MRI data is described in Figure 1.

**Figure 1.**
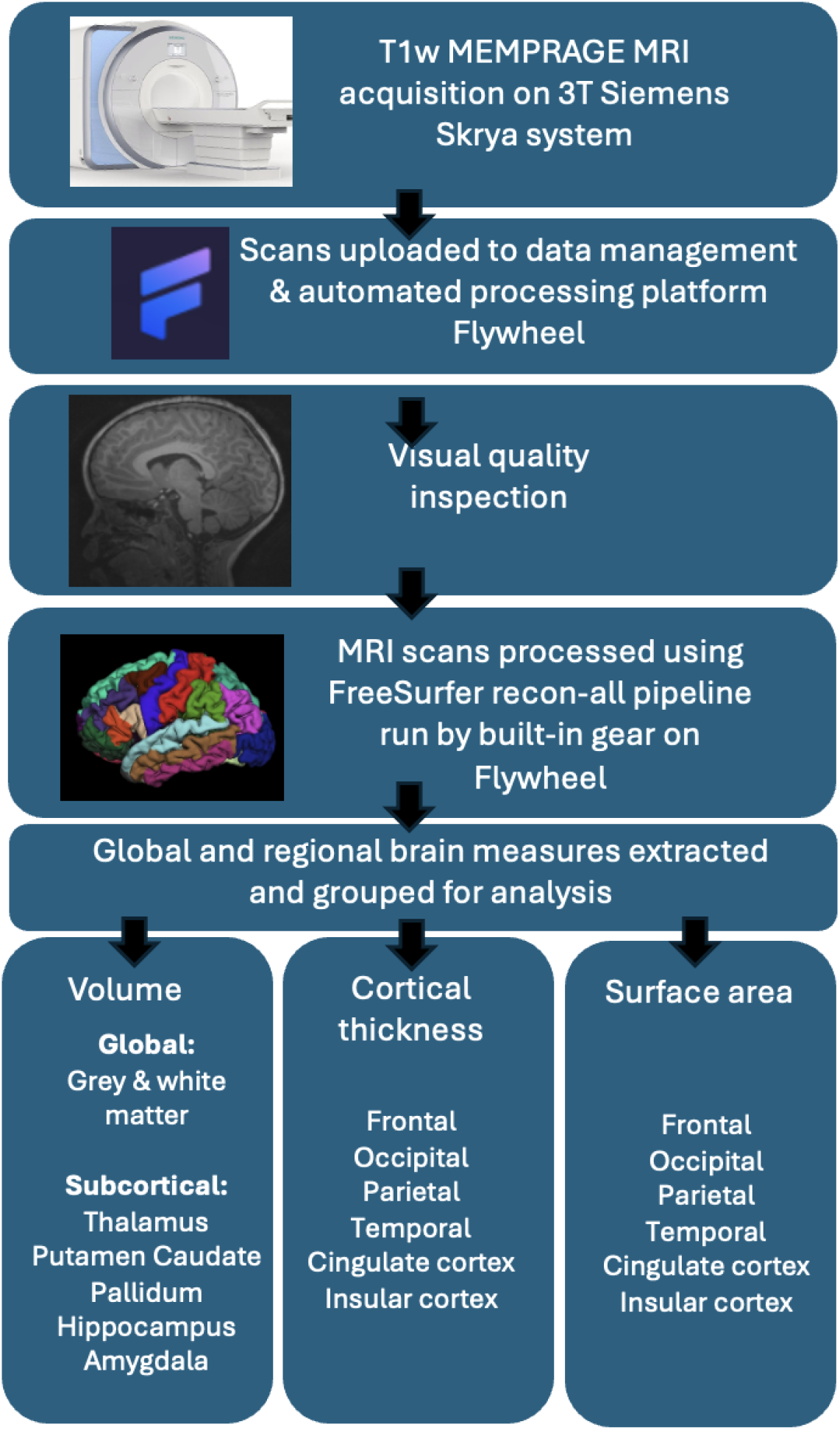
Workflow for acquisition, processing, and analysis of structural MRI data in the DolPHIN-2 PLUS neuroimaging sub-study High-resolution T1-weighted MPRAGE scans were acquired on a 3T Siemens Skyra system. Scans were uploaded to the Flywheel platform for centralized data management and automated preprocessing. After visual quality inspection, image processing was conducted on Flywheel using the built-in recon-all FreeSurfer (v7.2.0) pipeline. Global and regional brain metrics (volume, cortical thickness, and surface area) were extracted and grouped by lobe or structure using the Desikan-Killiany atlas. Regions of interest (ROIs) were selected a priori based on prior evidence from HIV-exposure studies and included both global and subcortical measures.

### Statistical Analysis

Sociodemographic and clinical were compared between groups (CHEU vs. CHU and DTG vs. EFV). For normally distributed continuous variables (e.g. age at scan, intracranial volume and maternal mean viral load [log-transformed]), comparisons were conducted using independent samples t-tests. The Mann-Whitney U test was applied to non-normally distributed continuous variables. Categorical variables were analyzed using chi-squared and Fisher’s exact tests, as appropriate.

Global and subcortical brain regions were selected a priori based on previously reported associations in CHEU. Given the limited existing literature on the effects of specific ART regimens on brain development, an exploratory approach was adopted for cortical thickness and surface area. Associations between exposure status (HIV and ART exposure) with brain structure (brain volume, cortical thickness and surface area) were examined using multiple linear regression models. To reduce the number of comparisons, bilateral subcortical ROI volumes were averaged. For cortical thickness and surface area, regions were grouped anatomically according to the Desikan-Killiany atlas into frontal, temporal, parietal, occipital, cingulate, and insular lobes^24^. All models were adjusted for total intracranial volume, sex, and age at scan. Analyses comparing DTG and EFV exposure additionally adjusted for maternal CD4 count.

In secondary analyses, associations between maternal HIV disease severity and child brain morphology were examined. Maternal viral load (measured during pregnancy and at delivery) and CD4 (measured during pregnancy) were evaluated as predictors of brain volume using linear regression. Cohen’s *d* effect size was estimated to assess the exposure and treatment effects and categorized according to conventional benchmarks^25^. Model residuals were assessed graphically to confirm assumptions of normality. All analyses were performed in STATA version 18.0 (StataCorp, College Station, TX), using a two-sided significance threshold of p < 0.05.

## Results

A total of 60 children successfully underwent T1w MRI scans, of which 2 were excluded due to the presence of severe motion artifact on visual quality inspection. The final sample included 58 children (30 male, 28 female) with high resolution T1w images. Of these, 25 were CHEU (13 male, 12 female, mean age 45.5 months) and 33 were CHU (17 male, 16 female, mean age 47 months). Within the CHEU group, 13 children were DTG-exposed (7 male, 6 female) and 12 were EFV-exposed (6 male, 6 female).

As shown in Table 1, demographic characteristics including age at scan, sex, maternal education, maternal employment, maternal substance use during pregnancy and maternal depression were similar between DTG- and EFV-exposed groups, and between CHEU and CHU. Maternal CD4 count at enrolment (prior to ART initiation) was significantly higher in the DTG group (median 520 cells/mm^3^, IQR 427-797 cells/mm^3^) compared to the EFV group (median 358.5 cells/mm^3^, IQR 289-447.5 cells/mm^3^, *p* = 0.039). Maternal viral load at enrolment and at delivery did not differ significantly between treatment groups (*p* = 0.480 and *p* = 0.378, respectively). At delivery, 11 (84.6%) mothers in the DTG group and 8 (66.7%) in the EFV group were virally suppressed (viral load < 50 copies/mL)

**Table 1.** Socio-demographic and clinical characteristics of children according to HIV exposure (CHU vs CHEU) and ART exposure (DTG vs. EFV) status.

| Variable | Total<br>(N = 58) | CHEU<br>(N = 25) | CHU<br>(N = 33) | p-value | DTG<br>(N = 13) | EFV<br>(N = 12) | p-value |
| --- | --- | --- | --- | --- | --- | --- | --- |
| *Sociodemographic characteristics* |  |  |  |  |  |  |  |
| Child age at scan<br>Mean (SD) (months) | 46.35 (3.97) | 45.50 (3.16) | 47.00<br>(4.42) | 0.133 | 45.69 (3.86) | 45.25 (2.34) | 0.730 |
| Sex |  |  |  |  |  |  |  |
| Male | 30 (51.7%) | 13 (52.0%) | 17<br>(51.5%) | 0.971 | 7 (53.8%) | 6 (50%) | 0.848 |
| Female | 28 (48.3%) | 12 (48.0%) | 16<br>(48.5%) |  | 6 (46.2%) | 6 (50%) |  |
| Maternal Education (Completed<br>secondary school) | 23 (39.7%) | 9 (36.0%) | 14<br>(42.4%) | 0.620 | 3 (23.1%) | 6 (50%) | 0.226 |
| Maternal Employment Status<br>(Employed) | 21 (36.2%) | 12 (48.0%) | 9 (27.3%) | 0.104 | 5 (38.5%) | 7 (58.3%) | 0.320 |
| Maternal alcohol use during<br>pregnancy | 19 (32.8%) | 8 (32.0%) | 11<br>(33.3%) | 0.915 | 2 (15.4%) | 6 (50%) | 0.097 |
| Maternal Depression | 8 (13.8%) | 2 (8.0%) | 6 (18.2%) | 0.445 | 1 (7.7%) | 1 (8.3%) | 1.000 |
| *Neuroanatomical variables* |  |  |  |  |  |  |  |
| Total intracranial volume (cm<sup>3</sup>) | 1291.97<br>(154.23) | 1286.62<br>(162.04) | 1296.02<br>(150.46) | 0.822 | 1279.42<br>(204.08) | 1294.42<br>(108.28) | 0.819 |
| *Maternal HIV variables in pregnancy* |  |  |  |  |  |  |  |
| Maternal CD4 count, median<br>(IQR) (cells/mm<sup>3</sup>) |  | 440 (346.0-660.0) |  |  | 520 (427.0-797.0) | 358.5 (289.0<br>-447.5) | 0.039 |
| ≤500 cells/mm<sup>3</sup> |  | 15 (60%) |  |  | 5 (38.5%) | 10 (83.3%) | 0.041* |
| >500 cells/mm<sup>3</sup> |  | 10 (40%) |  |  | 8 (61.5%) | 2 (16.7%) |  |
| Maternal Viral Load at<br>enrollment (copies/ml) |  |  |  |  |  |  |  |
| <50 copies/ml |  | 2 (8%) |  |  | 2 (15.4%) | 0 (0%) | 0.480 |
| ≥50 copies/ml |  | 23 (92%) |  |  | 11 (84.6%) | 12 (100%) |  |
| Maternal Viral Load at delivery<br>(copies/ml) |  |  |  |  |  |  |  |
| <50 copies/ml |  | 19 (76%) |  |  | 11 (84.6%) | 8 (66.7%) | 0.378 |
| ≥50 copies/ml |  | 6 (24%) |  |  | 2 (15.4%) | 4 (33.3%) |  |

Data are N (%), mean (SD), or median (IQR). Continuous variables were compared with unpaired t-tests if normally distributed or Mann-Whitney U tests if distribution was not normal; categorical variables were compared with chi-squared tests. \*p<0.05. Abbreviations: CHEU, children who are HIV-exposed uninfected; CHU, children who are HIV-unexposed; DTG, dolutegravir; EFV, efavirenz

### ART exposure and brain structure

Total intracranial volume was similar between those children exposed to DTG and EFV (1279 vs. 1294 cm^3^, *p*=0.819) (Table 1). Within the CHEU group, there were no differences in the brain volumes between DTG- and EFV-exposed children. This included total grey matter (adjusted difference -1386.54 mm^3^, Cohen’s *d* = -0.08 [95% CI -0.86 to 0.71], *p*=0.861) and white matter volume (9182.01 mm^3^, *d =* 0.66 [-0.16 to 1.46], *p* = 0.158). No significant differences were observed in subcortical ROIs, including the thalamus (-238.98 mm^3^, *d =* -0.32 [-1.12 to 0.47], *p* = 0.479), caudate (284.71 mm^3^, *d =* 0.32 [-0.48 to 1.10], *p*=0.484), putamen (714.46 mm^3^, *d* = 0.51 [-0.30 to 1.3], *p* = 0.272), pallidum (161.04 mm^3^, *d* = 0.48 [-0.32 to 1.27], *p* = 0.292), hippocampus (-37.96 mm^3^, *d =* -0.07 [-0.85 to 0,72], *p* = 0.885), and amygdala (-58.68 mm^3^, *d =* -0.20 [-0.98 to 0.59], *p* = 0.666) (Table 2). There were also no significant differences in cortical thickness or surface are between ART exposure (*p* > 0.1, Tables S1 – S2). Effect size estimates indicated a medium effect for the putamen (0.5 ≤ *d* < 0.8), small effects in the amygdala, thalamus, caudate and pallidum (0.2 ≤ *d* < 0.5), and very small effects in the hippocampus (*d* < 0.2) (Figure 2).

**Table 2.** Adjusted mean differences in global and regional grey matter volumes between children according to HIV exposure (CHEU vs CHU) and ART exposure (DTG vs. EFV) status.

| Brain Volume (mm<sup>3</sup>) | CHEU mean (SD) | CHU mean (SD) | Adjusted<sup>a</sup> difference (95% CI) | P-value | Cohen's d (Effect size) | DTG mean (SD) | EFV mean (SD) | Adjusted<sup>b</sup> difference (95% CI) | P-value | Cohen's d (Effect size) |
| --- | --- | --- | --- | --- | --- | --- | --- | --- | --- | --- |
| *Global* |  |  |  |  |  |  |  |  |  |  |
| Total Grey Matter | 671708.64 (52399.09) | 671498.51 (61727.25) | 1982.61 (-11855.57 to 15820.79) | 0.775 | 0.08 (-0.44 to 0.60) | 668574.10 (69326.79) | 675104.39 (26892.74) | -1386.54 (-17689.1 to 14916.02) | 0.861 | -0.08 (-0.86 to 0.71) |
| Cerebral White Matter | 345040.68 (29875.13) | 346601.48 (47679.93) | 3048.24 (-12903.24 to 18999.72) | 0.703 | 0.10 (-0.42 to 0.62) | 348985.69 (37365.59) | 340766.92 (19644.13) | 9182.01 (-3884.93 to 22248.95) | 0.158 | 0.66 (-0.16 to 1.46) |
| *Subcortical* |  |  |  |  |  |  |  |  |  |  |
| Thalamus | 12496.71 (761.45) | 12464.37 (1386.44) | 93.09 (-470.04 to 656.2) | 0.742 | 0.09 (-0.43 to 0.61) | 12393.98 (837.98) | 12608.01 (687.76) | -238.98 (-930.95 to 452.99) | 0.479 | -0.32 (-1.12 to 0.47) |
| Caudate | 7117.14 (948.43) | 7340.02 (1044.21) | -168.42(-657.99 to 321.16) | 0.493 | -0.19 (-0.71 to 0.34) | 7296.18 (1109.74) | 6923.12 (734.99) | 284.71 (-550.69 to 1120.09) | 0.484 | 0.32 (-0.48 to 1.10) |
| Putamen | 9111.72 (1576.99) | 9492.98 (1129.32) | -283.68 (-917.04 to 349.68) | 0.373 | -0.24 (-0.76 to 0.28) | 9447.34 (1446.21) | 8748.14 (1693.21) | 714.4637 (-606.60 to 2035.53) | 0.272 | 0.51 (-0.30 to 1.30) |
| Pallidum | 3393.42 (435.90) | 3515.10 (520.49) | -65.41 (-269.31 to 138.50) | 0.523 | -0.17 (-0.69 to 0.35) | 3483.33 (462.22) | 3296.03 (401.98) | 161.04 (-150.18 to 472.25) | 0.292 | 0.48 (-0.32 to 1.27) |
| Hippocampus | 6352.51 (805.88) | 6600.43 (842.71) | -189.21 (-511.06 to 132.64) | 0.244 | -0.32 (-0.84 to 0.21) | 6352.37 (989.86) | 6352.67 (589.99) | -37.96 (-581.77 to 505.85) | 0.885 | -0.07 (-0.85 to 0.72) |
| Amygdala | 2489.11 (495.79) | 2543.26 (336.75) | 35.31 (-186.29 to 115.67) | 0.641 | -0.13 (-0.65 to 0.39) | 2458.76 (626.37) | 2521.99 (325.64) | -58.68 (-339.14 to 221.79) | 0.666 | -0.20 (-0.98 to 0.59) |

Multiple linear regression estimates for HIV and ART exposure on global and subcortical brain volume. <sup>a</sup> Adjusted for age, sex, intracranial volume. <sup>b</sup> Adjusted for age, sex intracranial volume and maternal CD4. Global and subcortical volume (mean total of left and right hemispheres), mean differences (adjusted regression coefficients with 95% confidence intervals in multiple regression models), p-values, Cohen's D effect sizes (95% confidence intervals) are presented here. *Abbreviations: CHEU, Children who are HIV-exposed uninfected; CHU, children who are HIV-unexposed; DTG, dolutegravir; EFV, efavirenz; CI, confidence interval; SD, standard deviation*

**Figure 2.**
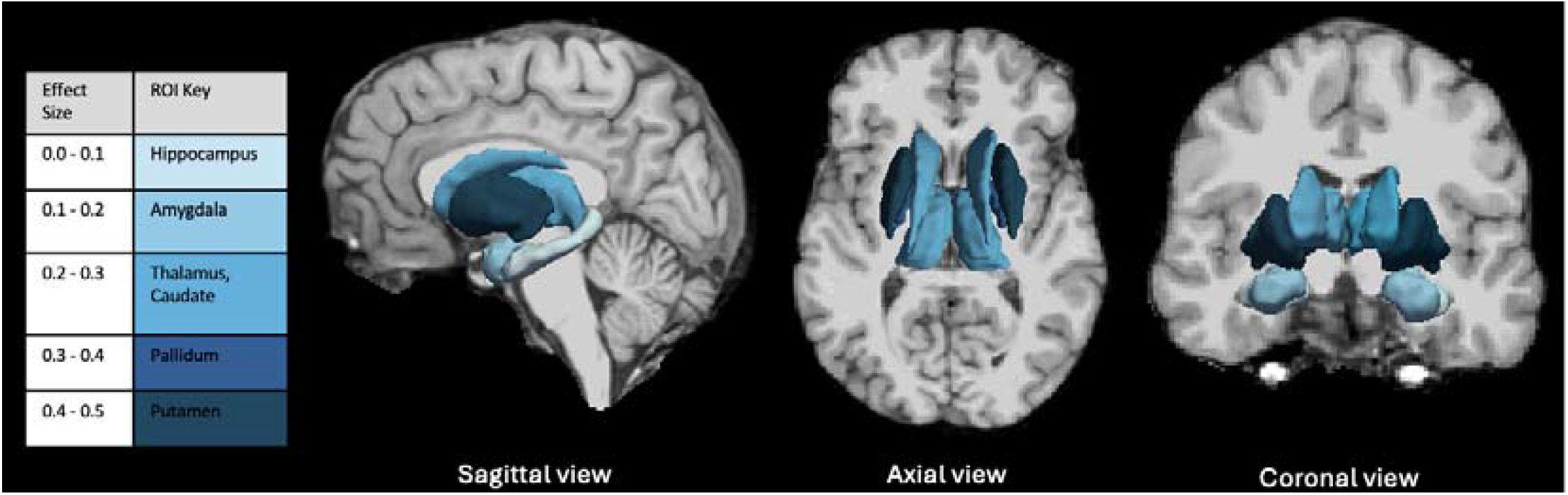
Schematic of the effect sizes of subcortical brain volumes differences in CHEU exposed to DTG versus EFV. The key represents the color coding of absolute effect sizes corresponding to brain volumes in subcortical regions of interest. Schematic created using Freesurfer version 7.2.0. *Abbreviations: ROI, region of interest; CHEU, children who are HIV-exposed uninfected; DTG, dolutegravir; EFV, efavirenz*

### HIV exposure and brain structure

Total intracranial volume was similar between CHEU and CHU (1286 vs.1296 cm^3^, *p* = 0.822) (Table 1). Multiple linear regression estimates showed no brain volume differences of CHEU in comparison to CHU in total grey matter (adjusted difference 1982.61 mm^3^, Cohen’s *d =* 0.08 [95% CI -0.44 to 0.60, *p* = 0.775) and total white matter (3048.24 mm^3^, *d =* -0.10 [-0.44 to 0.60], *p* = 0.775), or subcortical ROIs including the thalamus (93.09 mm^3^, *d =* 0.09 [-0.43 to 0.61], *p* = 0.742), caudate (-168.42 mm^3^, *d =* -0.19 [-0.71 to 0.34], *p* = 0.493 ), putamen (-283.68 mm^3^, *d =* -0.24 [-0.76 to 0.28], *p* = 0.373 ), pallidum (-65.41 mm^3^, *d =* - 0.17 [-0.69 to 0.35], *p* = 0.523 ), hippocampus (-189.21 mm^3^, *d =* -0.32 [-0.84 to 0.21], *p* = 0.244 ), and amygdala (35.31 mm^3^, *d =* -0.13 [-0.65 to 0,39], *p* = 0.641 ) (Table 2). Similarly, multiple linear regression estimates showed no association of HIV exposure with brain surface area or cortical thickness (*p* > 0.05, Tables S1 S2).

### Maternal HIV disease severity and child brain structure

In exploratory analyses, no significant associations were observed between maternal CD4 count during pregnancy, analyzed as both continuous and dichotomized variables, and global or regional brain volumes in children (*p* > 0.3, Table S3). Similarly, maternal viral load (log-transformed), measured at enrolment in the third trimester prior to ART initiation and at delivery, was no associated with child brain volume outcomes (*p* > 0.1, Table S4)

## Discussion

Dolutegravir is established as the first-line treatment for HIV in pregnancy, yet data on its long-term impact on child neurodevelopment remain limited. In this study, we found no differences in brain morphology, including volume, cortical thickness, or surface area, between children with in-utero exposure to DTG versus EFV in late-stage pregnancy, nor between CHEU and CHU. To our knowledge, this is the first study to use MRI to assess brain structure of children with perinatal exposure to DTG. These findings contribute novel neuroimaging data and insights into neurodevelopment in the context of this more recent treatment.

Children with in-utero exposure to DTG- and EFV-based ART initiated in late pregnancy demonstrated comparable brain structure across all measured domains. A key advantage of DTG over EFV in pregnancy is its superior efficacy in achieving maternal viral suppression at the time of birth. The DolPHIN-2 clinical trial demonstrated pregnant women who initiated DTG-based therapy in the third trimester were more likely to achieve viral suppression (<50 copies/mL) at delivery ^20^. In this neuroimaging sub study, conducted with a smaller South African subset of participants from DolPHIN-2, the rate of viral load suppression at the time of delivery were similar between ART groups. This comparable viral suppression in this sub sample may partly explain the lack of observed differences in brain structure between DTG- and EFV-exposed children. Early and effective viral suppression of HIV is critical for several general health reasons such as preventing HIV transmission but also for reducing the potential neurotoxic effects of the virus and associated maternal immune activation on fetal brain development. Additionally, high levels of alcohol exposure, which were observed across all groups in this study, may have obscured more subtle ART or HIV-related effects on neurodevelopment. To our knowledge, only one other neuroimaging study has been set up to investigate brain structure in CHEU with a focus on ART exposure. In that study, neonates with prenatal exposure to HIV and EFV-based ART had smaller left putamen and bilateral caudate volumes compares to their HIV-unexposed counterparts^17^. Importantly, reductions in caudate volume were dependent on the duration of in-utero ART-exposure, with effects only seen in neonates born to mothers initiating ART post-conception^17^. This suggests that earlier and sustained ART may offer neuroprotection to subcortical brain structures. Although our study could not assess the impact of ART initiation timing, this remains as an important consideration in optimizing fetal brain development among CHEU, particularly in the context of expanding DTG use in pregnancy.

We conducted an exploratory analysis within the CHEU group to examine whether maternal HIV disease severity, as measured by viral load and CD4 count, was associated with child brain volume. No significant associations were observed in our cohort. Maternal immune health during pregnancy is thought to play and important role in early development as previous studies have shown that lower maternal CD4 and higher viral load are associated with an increased risk of infant morbidity and mortality^26^. Although the exact mechanisms underlying this remain unclear, they are likely multifactorial^27^. Maternal immune dysregulation has previously been linked to various neurological disorders and abnormal brain development^28^. It is therefore plausible that chronic exposure to maternal inflammation caused by HIV infection may impact fetal brain development with lasting effects. This hypothesis is supported by findings from the Drakenstein Child Health Study (DCHS) birth cohort, where lower maternal CD4 count was found to be associated with smaller grey matter and putamen volumes in CHEU as neonates and 2-3 years of age respectively^18,19,29^. While both the DCHS and DolPHIN-2 PLUS studies were conducted in similar settings in South

Africa, the DolPHIN-2 PLUS children represent a more recent cohort of children born between 2018 and 2019. Global and national efforts to expand ART coverage and strengthen prenatal care in mothers living with HIV in South Africa, may be contributing to improved maternal health and pregnancy outcomes, and consequently healthier children^30,31^. Although our analyses were limited by small sample size, it is possible the lack of association between maternal CD4 and child brain volume reflects better overall prenatal health and HIV management in this more recent cohort. Further research is needed to determine if this is evident in other current birth cohorts.

When comparing CHEU and CHU groups, we found no associations between HIV exposure and brain and brain volume, surface area or thickness. This contrasts previous studies which have reported altered structural brain development in CHEU, however none have examined children with DTG exposure ^17-19^. Recent MRI research conducted in South Africa have reported reduced volumes in grey matter, and basal ganglia regions including the caudate and left putamen, in CHEU when compared to their unexposed counterparts^17,18,32,33^. In addition to altered brain volume, increased cortical thickness in prefrontal regions such as the medial orbitofrontal cortex, have been found in CHEU at 2-3 years of age^34^. The similar brain structure observed between CHEU and CHU in this study may have been driven by the use of newer and more effective HIV treatments for use in late pregnancy, which may be protecting the developing brain from the HIV-related effects seen in older studies. Additionally, mothers in the CHEU group were part of the DolPHIN-2 clinical trial, as a result, may have had better access to perinatal and follow-up care and support which could have mitigated the neurodevelopmental impact associated with exposure to HIV and ART in early life. This study had a smaller sample size compared to DCHS and therefore may have been underpowered to detect the subtle effects of HIV exposure on the developing brain. It is also possible that other unmeasured confounders, such as maternal health, quality of prenatal care and environmental exposures present in the CHU group, may have influenced brain development and contributed to the observed lack of differences between CHEU and CHU.

A key strength of this study was the availability of detailed maternal HIV health metrics gathered in mothers of CHEU in late pregnancy, which allowed us an in-depth analysis of maternal HIV and ART and brain structure in CHEU. However, the timing of maternal HIV diagnosis and infection prior to trial enrolment was not available, limiting our ability to determine ART duration or whether mothers in the CHEU group had chronic HIV infection or were recently seroconverted. Consequently, immune status prior to the third trimester could not be assessed. Additionally, the pregnancy health records of mothers of CHU were not available as this group was recruited postnatally, thus limiting our ability to assess other important factors related to maternal immune status that may impact brain development regardless of HIV/ART exposure. Other limitations inherent to paediatric MRI studies may result from scanning young children in non-sedated sleep. Our small sample size, although moderate for an MRI study, reduces the statistical power and generalizability of these findings and may have limited our ability to detect between-group differences in brain structure. We acknowledge there may have been other unmeasured factors which could have impacted our findings. Nonetheless, these findings contribute valuable insight into the relationship between maternal ART and neurodevelopment. Future research is needed to investigate the developmental trajectory of children exposed to DTG and other newer ART, the effects of ART timing and duration, and the relationship between maternal immune health and brain structure. Larger sample sizes assessing both structural and functional outcomes will further clarify the long-term impact of DTG on child neurodevelopment.

## Conclusion

Overall, children exposed to DTG or EFV in late pregnancy have similar brain structure in early life to children who were not exposed to HIV or ART. However, more research with larger sample sizes is needed to replicate these findings and elucidate any protective effect that earlier ART initiation may offer. Our findings provisionally support the use of DTG in late pregnancy with respect to child neurodevelopmental outcomes. Ongoing monitoring and further research are crucial to fully elucidate the effects of prenatal exposure to antiretroviral therapies on long-term neurodevelopment. These efforts are essential for optimizing treatment guidelines and improving outcomes for children born to mothers living with HIV

## Data Availability

All data produced in the present study are available upon reasonable request to the authors

## Notes

## Acknowledgments

We acknowledge and thank the children and families of the DolPHIN-2 PLUS study, the contributions of study staff as well as the radiographers at Cape Universities Body Imaging Centre who made this work possible.

## Author Contributions

Layla E Bradford: Conceptualization, methodology, software, analysis, investigation, data curation, manuscript writing, manuscript review and editing, project administration

Jessica E Ringshaw: Methodology, software, investigation, data curation, manuscript review and editing, project administration

Catherine J Wedderburn: Methodology, software, analysis, manuscript writing, manuscript review and editing, supervision

Kirsten A Donald: Conceptualization, methodology, analysis, manuscript writing, manuscript review and editing, supervision, project administration

Lauren Davel: Investigation, data curation, manuscript review and editing, project administration

Catherine Orrell: Methodology, manuscript review and editing

Niall J Bourke: Methodology, software, manuscript review and editing Steve CR Williams: Methodology, software, manuscript review and editing

Landon Myer: Conceptualization, methodology, manuscript review and editing, funding acquisition

Thokozile R Malaba: Conceptualization, methodology, manuscript review and editing, project administration

Helene Theunissen: Data curation, manuscript review and editing, project administration Catriona Waitt: Conceptualization, manuscript review and editing

Helen Reynolds: Manuscript review and editing, project administration

Saye Khoo: Conceptualization, methodology, manuscript review and editing, funding acquisition

Miriam Taegtmeyer: Conceptualization, manuscript review and editing Nengjie He: Manuscript review and editing

Jim Read: Data curation, manuscript review and editing Lucy Read: Data curation, manuscript review and editing Duolao Wang: Methodology, manuscript review and editing

Angela Colbers: Methodology, manuscript review and editing

