## Supplementary Table S1 for "Brain structure of South African children born to mothers on dolutegravir versus efavirenz-based ART"

**Supplementary Table S1.** Adjusted mean differences of brain surface areas between children according to HIV exposure (CHEU vs CHU) and ART exposure (DTG vs. EFV) status

| **Total Surface Area** | **CHEU mean (SD)** | **CHU mean (SD)** | **Adjusted^a^ difference** | **P-value** | **Cohen’s d (effect size)** | **DTG**  **mean (SD)** | **EFV**  **mean (SD)** | **Adjusted^b^ difference** | **P-value** | **Cohen’s d (effect size)** |
| --- | --- | --- | --- | --- | --- | --- | --- | --- | --- | --- |
| Frontal Lobe | 55607.56 (5343.82) | 55898.91 (6059.79) | 83.84 (-1995.61 to 2163.3) | 0.936 | 0.02 (-0.50 to 0.54) | 55528.62 (7086.20) | 55693.08 (2740.52) | 480.35 (-2522.23 to 3482.93) | 0.741 | 0.15 (-0.64 to 0.93) |
| Temporal Lobe | 29873.16 (2880.82) | 29972.03 (3378.98) | 84.42 (-1006.27 to 1175.12) | 0.877 | 0.04 (-0.48 to 0.56) | 30065.69 (3871.58) | 29664.58 (1290.02) | 580.22 (-917.13 to 2077.56) | 0.427 | 0.36 (-0.43 to 1.12) |
| Parietal Lobe | 44070.68 (4236.88) | 44114.15 (5804.56) | 359.00 (-1883.95 to 2601.5) | 0.749 | 0.09 (-0.43 to 0.61) | 44665.23 (5117.48) | 43426.58 (3118.72) | 830.43 (-2429.05 to 4089.92) | 0.600 | 0.24 (-0.55 to 1.022) |
| Occipital Lobe | 22500.32 (3294.56) | 22089.39 (2786.22) | 341.89 (-1109.17 to 1792.94) | 0.638 | 0.13 (-0.39 to 0.65) | 21674.62 (3578.02) | 23394.83 (2834.96) | -1384.77 (-3659.57 to 890.02) | 0.218 | -0.57 (-1.36 to 0.24) |
| Cingulate Cortex | 6757.32 (692.92) | 6765.67 (898.98) | 39.97 (-294.94 to 374.88) | 0.812 | 0.06 (-0.46 to 0.58) | 6839.08 (883.10) | 6668.75 (424.69) | 166.01 (-360.32 to 692.34) | 0.517 | 0.29 (-0.50 to 1.08) |
| Insular Cortex | 4217.88 (393.99) | 4108.46 (495.74) | 111.76 (-120.30 to 343.82) | 0.338 | 0.26 (-0.26 to 0.78) | 4240.39 (381.40) | 4193.50 (422.78) | 97.26 (-244.00 to 438.51) | 0.558 | 0.27 (-0.53 to 1.05) |

Multiple linear regression estimates for HIV and ART exposure on brain surface area.^a^ Adjusted for age, sex, intracranial volume. ^b^Adjusted for age, sex intracranial volume and maternal CD4. Total surface area (total mean of left and right hemispheres combined), mean differences (adjusted regression coefficients with 95% confidence intervals in multiple regression models), p-values are presented here. *Abbreviations: CHEU, Children who are HIV-exposed uninfected; CHU, children who are HIV-unexposed; DTG, dolutegravir; EFV, efavirenz; CI, confidence interval; SD, standard deviation.*
