## Supplementary Table S2 for "Brain structure of South African children born to mothers on dolutegravir versus efavirenz-based ART"

**Supplementary Table S2.** Adjusted mean differences of cortical thickness between children according to HIV exposure (CHEU vs CHU) and ART exposure (DTG vs. EFV)

| **Mean Cortical Thickness** | **CHEU**  **mean (SD)** | **CHU**  **mean (SD)** | **Adjusted^a^ difference** | **P-value** | **Cohen’s d**  **(effect size)** | **DTG**  **mean (SD)** | **EFV**  **mean (SD)** | **Adjusted^b^ difference** | **P-value** | | **Cohen’s d (effect size)** |
| --- | --- | --- | --- | --- | --- | --- | --- | --- | --- | --- | --- |
| Frontal Lobe | 2.89 (0.11) | 2.91 (0.14) | -0.03 (-0.10 to 0.04) | 0.406 | -0.23 (-0.75 to 0.30) | 2.90 (0.13) | 2.88  (0.10) | 0.02 (-0.09 to 0.12) | 0.776 | 0.13 (-0.66 to 0.91) | |
| Temporal Lobe | 2.95 (0.10) | 2.94 (0.14) | -0.01 (-0.08 to 0.06) | 0.750 | -0.09 (-0.61 to 0.43) | 2.94 (0.09) | 2.96  (0.11) | -0.02 (-0.12 to 0.08) | 0.656 | -0.20 (-0.98 to 0.59) | |
| Parietal Lobe | 2.70 (0.08) | 2.69 (0.11) | 0.01 (-0.04 to 0.06) | 0.735 | 0.01 (-0.04 to 0.06) | 2.71 (0.09) | 2.70  (0.08) | 0.02 (-0.05 to 0.10) | 0.538 | 0.28 (-0.52 to 1.06) | |
| Occipital Lobe | 2.24 (0.10) | 2.24 (0.11) | 0.01 (-0.05 to 0.06) | 0.861 | 0.05 (-0.47 to 0.57) | 2.27 (0.10) | 2.22  (0.10) | 0.07 (-0.02 to 0.17) | 0.112 | 0.73 (-0.09 to 1.54) | |
| Cingulate Cortex | 2.71 (0.14) | 2.69 (0.15) | 0.00 (-0.07 to 0.07) | 0.977 | -0.01 (-0.53 to 0.51) | 2.70 (0.15) | 2.73  (0.14) | -0.04 (-0.16 to 0.09) | 0.575 | -0.25 (-1.04 to 0.54) | |
| Insular Cortex | 3.14 (0.15) | 3.21 (0.19) | -0.09 (-0.18 to 0.00) | 0.052 | -0.54 (-1.06 to -0.01) | 3.13 (0.12) | 3.16  (0.18) | -0.06 (-0.20 to 0.09) | 0.412 | -0.37 (-1.16 to 0.43) | |

Multiple linear regression estimates for HIV and ART exposure on brain cortical thickness..^a^ Adjusted for age and sex,. ^b^Adjusted for age, sex and maternal CD4. Cortical thickness (mean of left and right hemispheres), mean differences (adjusted regression coefficients with 95% confidence intervals in multiple regression models), p-values are presented here. *Abbreviations: CHEU, Children who are HIV-exposed uninfected; CHU, children who are HIV-unexposed; DTG, dolutegravir; EFV, efavirenz; CI, confidence interval; SD, standard deviation.*
