## Supplementary Table S3 for "Brain structure of South African children born to mothers on dolutegravir versus efavirenz-based ART"

**Supplementary Table S3.** Adjusted mean differences in global and regional grey matter volumes between children according to maternal CD4 count in pregnancy

| **Maternal CD4 count at enrolment (cells/mm3)** | **Mean (SD)** | **Adjusted^a^ coefficient (95% CI)** | **P-value** |
| --- | --- | --- | --- |
| *Global* |  |  |  |
| **Total Grey**  CHU  CD4 >500 cells/mm^3^  CD4 ≤500 cells/mm^3^ | 671498.50 (61727.25)  674992.20 (55579.73)  669519.60 (52040.02) | Reference  -3888.47 (-22476.46 to 14699.53)  6181.67 (-10266.23 to 22629.56) | 0.613 |
| **Cerebral White matter**  CHU  CD4 >500 cells/mm^3^  CD4 ≤500 cells/mm^3^ | 346601.50 (47679.93)  353921.30 (34011.71)  339120.30 (26315.35) | Reference  4589.18 (-17013.39 to 26191.75)  1946.14 (-17169.25 to 21061.53) | 0.909 |
| *Subcortical* |  |  |  |
| **Thalamus**  CHU  CD4 >500 cells/mm^3^  CD4 ≤500 cells/mm^3^ | 12464.37 (1386.44)  12461.77 (912.08)  12520.01 (676.55) | Reference  -76.46 (-836.13 to 683.20)  214.35 (-457.85 to 886.55) | 0.757 |
| **Caudate**  CHU  CD4 >500 cells/mm^3^  CD4 ≤500 cells/mm^3^ | 7340.02 (1044.21)  7278.99 (1091.83)  7009.25 (862.85) | Reference  -117.44 (-780.41 to 545.53)  -204.88 (-791.52 to 381.76) | 0.772 |
| **Putamen**  CHU  CD4 >500 cells/mm^3^  CD4 ≤500 cells/mm^3^ | 9492.98 (1129.32)  9531.81 (1211.55)  8831.67 (1763.39) | Reference  -41.76 (-893.91 to 810.39)  -456.70 (-1210.74 to 297.34) | 0.470 |
| **Pallidum**  CHU  CD4 >500 cells/mm^3^  CD4 ≤500 cells/mm^3^ | 3515.10 (520.49)  3599.47 (366.87)  3256.06 (434.35) | Reference  53.95 (-217.78 to 325.68)  -150.78 (-391.22 to 89.67) | 0.346 |
| **Hippocampus**  CHU  CD4 >500 cells/mm^3^  CD4 ≤500 cells/mm^3^ | 6600.43 (842.71)  6525.16 (1008.88)  6237.41 (650.79) | Reference  -139.60 (-575.18 to 295.97)  -224.69 (-610.11 to 160.74) | 0.481 |
| **Amygdala**  CHU  CD4 >500 cells/mm^3^  CD4 ≤500 cells/mm^3^ | 2543.26 (336.75)  2554.92 (466.18)  2445.24 (525.85) | Reference  -21.01 (-225.48 to 183.46)  -45.53 (-226.46 to 135.40) | 0.878 |

Multiple linear regression estimates for maternal CD4 and brain volume.^a^ Adjusted for age, sex and intracranial volume. Global and subcortical volume (mean total of left and right hemispheres) dichotomized by maternal CD4 count, mean differences (adjusted regression coefficients with 95% confidence intervals in multiple regression models), overall p-values are presented here. *Abbreviations: CHU, children who are HIV-unexposed; CI, confidence interval; SD, standard deviation.*
